# Inferring blood volume collected on plasma-separating dried blood spots using mass and image analysis

**DOI:** 10.64898/2025.11.28.25341240

**Authors:** Elise C. Dietmann, Michelle D. Stephens, Olivia Krebs, Everlyne N. Nkadori, Judy Wang, Bradon R. McDonald, Pallavi Tiwari, Stephanie M. McGregor, Muhammed Murtaza

**Affiliations:** Center for Precision Medicine, University of Wisconsin–Madison, Madison, WI, USA; Department of Surgery, School of Medicine and Public Health, University of Wisconsin–Madison, Madison, WI, USA; Carbone Cancer Center, University of Wisconsin–Madison, Madison, WI, USA; Departments of Radiology, Biomedical Engineering, and Medical Physics, University of Wisconsin–Madison, Madison, WI, USA; Department of Biomedical Engineering, Case Western Reserve University, Cleveland, OH, USA; Department of Pathology and Laboratory Medicine, School of Medicine and Public Health, University of Wisconsin-Madison, Madison, WI, USA

## Abstract

**Background:** Plasma-separating dried blood spots (psDBS) allow measurement of cancer biomarkers such as cell-free DNA concentration, enabling distributed sample collection, as well as ambient storage and shipment. However, unlike volumetric blood tube collections, blood volume collected using psDBS can vary between individuals. We evaluated whether analysis of psDBS mass, area, and color can be used to infer blood volume, and enable downstream measurement of circulating analyte concentrations.

**Methods:** Whole blood from healthy individuals and cancer patients was collected and used to prepare psDBS with volumes of 50 to 250 µL. Total area and mean grey value (MGV) of the erythrocyte region were measured using image segmentation. We developed a linear regression model to predict whole blood volume using mass, area, and MGV. DNA from psDBS was extracted, then quantified using a quantitative PCR assay targeting L1PA2.

**Results:** We analyzed 261 psDBS samples from 43 healthy individuals and 118 cancer patients. Using 110 samples from 10 healthy individuals, the linear regression model showed a strong correlation between actual and predicted blood volume (Pearson r = 0.97, RMSE = 5.1 µL). Using 151 samples from 33 separate healthy individuals and 118 samples from cancer patients, the model predicted blood volume with high accuracy (RMSE = 15.1 µL and 13.3 µL, respectively). Plasma DNA concentration in psDBS was moderately correlated with matched blood tubes (Spearman rho = 0.62).

**Conclusion:** Our results show estimating blood volume collected on psDBS is feasible using mass and image analysis. This approach enables calculation of circulating concentration for plasma analytes measured using psDBS.

## Background

The field of precision medicine continues to develop new blood-based assays for cancer detection and response monitoring during treatment. Recent work has investigated using biomarkers such as circulating DNA, RNA, or proteins from cancer cells, demonstrating their potential for early detection, prognostication, detection of minimal residual disease, and treatment response monitoring [1], [2]. In resource limited environments such as in rural areas, clinical implementation and access to blood-based assays is limited by the infrastructure required for phlebotomy, blood processing, and cold-chain storage and shipment. High costs associated with logistics of blood collection are especially prohibitive in low- and middle-income countries, where cancer-related death rates are disproportionately high despite lower incidence rates [3]. Thus, there is a need for more cost-effective and accessible blood sampling methods to reduce these care gaps [4].

One alternative to conventional phlebotomy is collection of dried blood spots (DBS). Recent studies suggest DBS samples may enable cancer detection and monitoring from a few drops of blood (∼150 µL) [5], [6]. DBS samples can be self-collected by fingerstick, stored at ambient temperature, and shipped inexpensively without the need for temperature control. More recently, plasma-separating DBS (psDBS) membranes have enabled collection of dried plasma samples to measure serum and plasma analytes while excluding background noise from other blood components [5]. Potential cancer biomarkers in plasma include cell-free DNA, proteins, cytokines and metabolites [2], [7]. psDBS are a promising, low-cost, and logistically simple alternative to phlebotomy that could improve access to blood-based early detection of cancer in resource-limited environments.

One barrier to further development and validation of cancer biomarkers in psDBS samples is the variability and inability to directly measure how much blood is collected in each sample. For most biomarkers, volume of blood or plasma collected or effectively analyzed is needed to calculate circulating concentrations, which are more clinically meaningful than eluant concentrations [8]. Unlike a blood tube where blood volume collected or volume of plasma analyzed downstream can be directly measured, volume of blood collected in psDBS samples can vary due to differences in sample collection methods such as operator experience, needle gauge and number of blood drops collected, physiology such as hydration levels, hematocrit or coagulation profiles, and external environmental conditions such as humidity and temperature [9].

To address this gap, we hypothesized that by measuring the psDBS mass, area of erythrocyte spread and brightness, we can estimate the volume of blood collected, and it can be used to calculate the circulating concentrations of plasma analytes. We developed a multiple linear regression model using psDBS features to infer collected blood volume. We found that by including multiple features into our model, we can estimate volume of blood collected in psDBS samples. Using these volumes to calculate circulating concentrations of plasma DNA, we observed a correlation between psDBS and corresponding blood tubes.

## Materials and methods

### Study design and cohort

The aim of this study was to develop a method of inferring collected blood volume from psDBS samples. In addition, we planned to use inferred blood volumes to calculate circulating concentration of plasma DNA in patients with cancer and healthy individuals, as an example of a plasma biomarker. On visual inspection, we observed physical and optical features that represent differences in total blood volume spotted on psDBS samples (Fig. 1a). Based on these observations, we hypothesized that by measuring the psDBS mass and erythrocyte area and brightness, we can estimate the volume of blood collected.

**Fig. 1:**
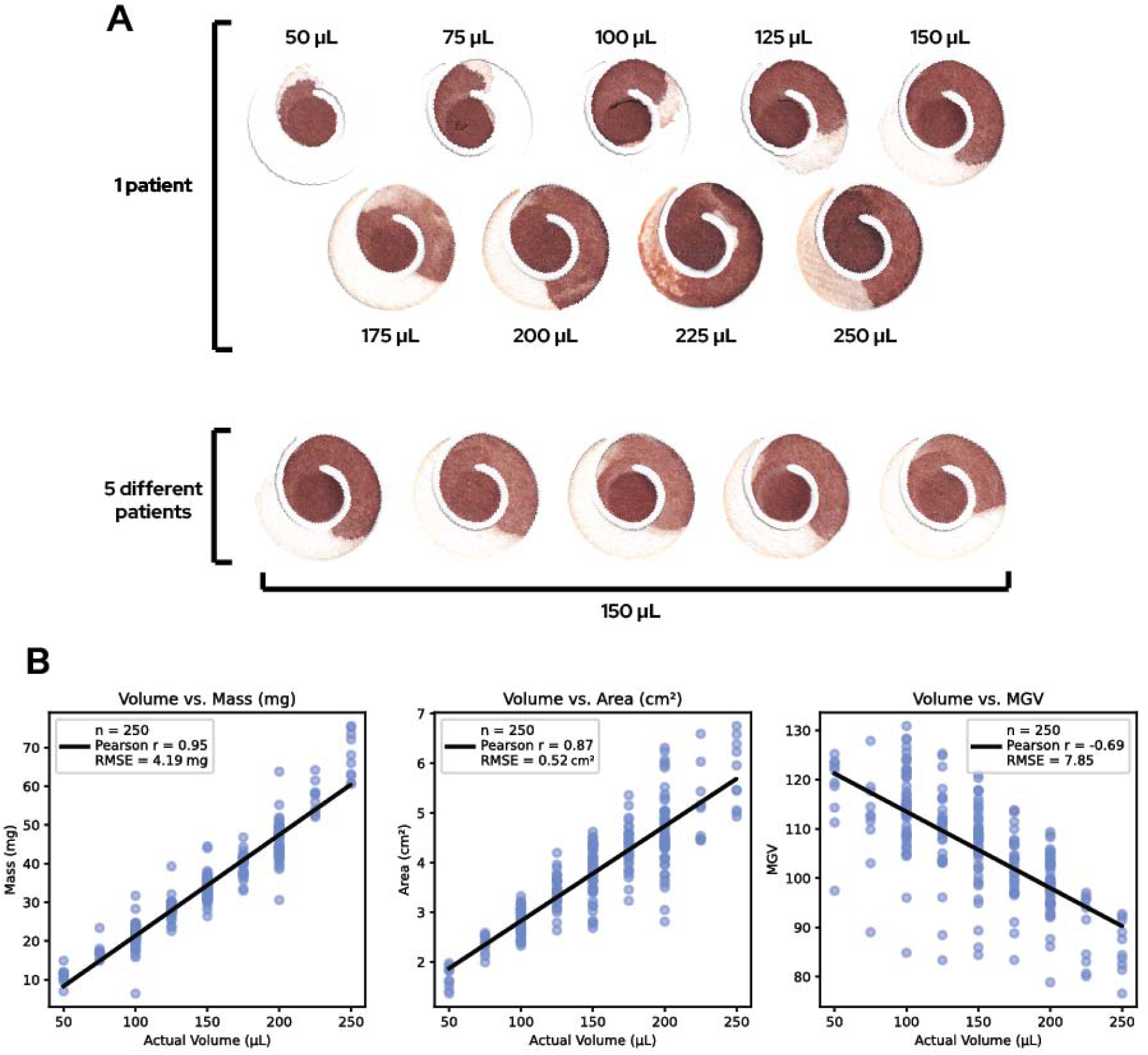
Evaluation of psDBS mass, blood spread area and intensity in HemaSpot samples. **(a)** Samples of different volumes from one patient have visible differences in the erythrocyte area and the brightness of the psDBS as volume increases. Samples of the same volume from five different patients have visible differences in the erythrocyte area and the brightness of the DBS due to biological variability. **(b)** There are positive correlations between the actual volume of the sample and mass and area (Pearson r = 0.95 and Pearson r = 0.87, respectively). There is a negative correlation between the actual volume of the sample and MGV (Pearson r = -0.69).

Overall, samples were obtained from two sources. First, we performed feature exploration using psDBS prepared from commercially obtained whole blood samples from healthy individuals (BioIVT, Westbury, NY). Second, we trained and validated our model for volume inference using psDBS prepared from whole blood samples obtained from healthy volunteers and cancer patients enrolled by the University of Wisconsin-Madison Carbone Cancer Center BioBank (UWCCC BioBank). Whole blood was collected under a protocol approved by the University of Wisconsin-Madison IRB (protocol number 2016-0934). Whole blood from patients with cancer, including head and neck, gynecologic and genitourinary, breast, and pancreatic cancers, was collected prior to surgery (Table 1).

**Table 1.** Description of the study cohort and participants.

|  | Source | Samples<br>n (%) | Individuals<br>n (%) | Age<br>median (range) | Sex<br>n (%) | Disease Status<br>n (%) |
| --- | --- | --- | --- | --- | --- | --- |
| Feature exploration | Commercial | 250 (100) | 30 (100) | 37.5 (18-66) | M: 15 (50)<br>F: 15 (50) | Healthy: 30 (100) |
| Model training | UWCCC<br>Biobank | 110 (100) | 10 (100) | NA | M: 2 (20)<br>F: 8 (80) | Healthy: 10 (100) |
| Model testing | UWCCC<br>Biobank | 151 (100) | 151 (100) | 61 (24-90) | M: 54 (35.8)<br>F: 97 (64.2) | Healthy: 33 (21.9)<br>Cancer:<br>Head and neck: 32 (21.2)<br>Gynecological: 25 (16.6)<br>Genitourinary: 20 (13.2)<br>Breast: 19 (12.6)<br>Pancreatic: 13 (8.6)<br>Other: 9 (5.9) |

### Sample collection and processing

Commercially obtained whole blood samples were collected in EDTA, shipped overnight to our lab at 4 °C, and psDBS samples were prepared at 50 to 250 µL volumes upon receipt. For patients and volunteers enrolled locally, whole blood was collected in EDTA and spotted within one hour by pipetting 50 to 250 µL onto the blood separation membrane of a psDBS collection device (HemaSpot™ SE, SpotOnSciences, San Francisco, CA). Blood on the membrane was fully dried before the device was stored at room temperature with a desiccant packet in a zippered plastic bag. An additional set of psDBS samples was prepared from locally collected samples using an alternative psDBS collection device (HemaSep™ Strip card, Ahlstrom, Helsinki, Finland) at 30 to 70 µL volumes, and used to evaluate generalizability of our approach.

The mass of each psDBS was measured using an ML54T/00 analytical balance (Mettler Toledo, Columbus, OH). Images were scanned in 24-bit color and 600 dots per inch (dpi) using a CanoScan LiDE 400 flatbed scanner (Canon, Huntington, NY). Each scan was automatically cropped in the software and was saved in JPG format (compressed). To measure and account for variation in scanner performance, a Delta 1 gray card (B&H Photo, New York, NY) was scanned before, during, and/or after scanning any psDBS (fig. S1).

### Image processing and feature extraction

Area and brightness of the psDBS were measured using an image segmentation approach (Fig. 2a). To measure the area of the psDBS, the total area of the erythrocyte region was extracted from the scan. Each psDBS scan underwent K-means clustering (K=2) to separate the erythrocyte region from the plasma region. A binary mask was created using Otsu’s thresholding, and the largest contour was assumed to correspond to the erythrocyte region. The total area of the erythrocytes was calculated in cm².

**Fig. 2:**
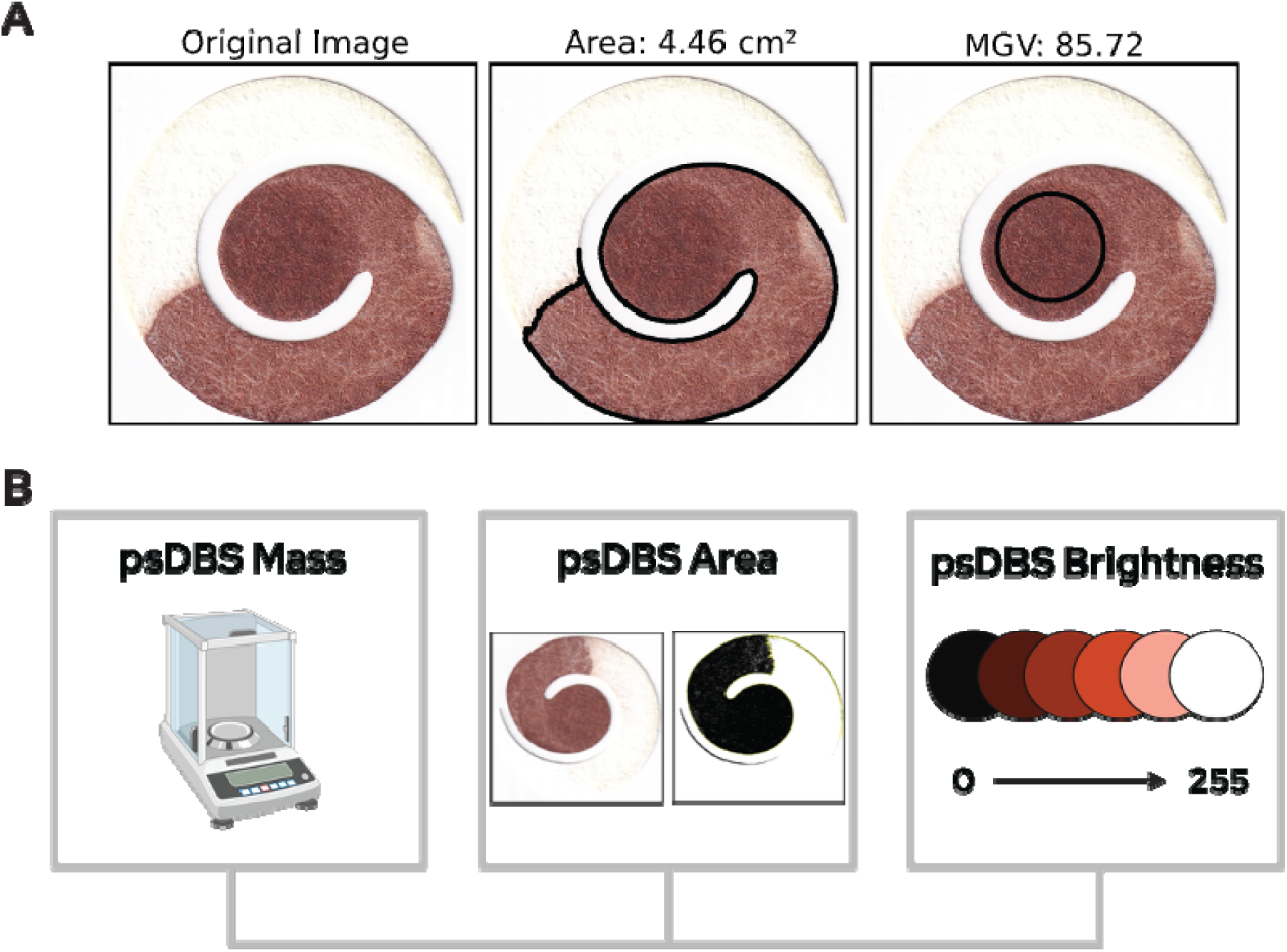
Image segmentation of Hemaspot samples and model development. **(a)** Area and brightness of the psDBS were measured using an image segmentation approach to quantitatively measure imaging features for use in the volume inference model. After K-means clustering (K=2) and the creation of a binary mask using Otsu’s thresholding, the largest contour was assumed to correspond to the erythrocyte region. The total area of the erythrocytes was calculated in cm² and used as the ‘area’ feature in the volume inference model. A mean gray value (MGV, scale 0-255) was calculated and used as the ‘brightness’ feature of the model. **(b)** The multiple linear regression model developed to predict whole blood volume uses the psDBS mass, erythrocyte area, and erythrocyte mean grey value as predictors.

To measure the brightness of the psDBS, a mean gray value (MGV) was calculated by averaging the pixel values across the three color channels in a region of interest centered within the erythrocyte area of the original image. Bright (near-white) pixels within the region that had an MGV value greater than 200 were excluded.

### Volume inference model development

Using the mass of the psDBS, erythrocyte area of the psDBS, and erythrocyte mean grey value of the psDBS as predictors, a multiple linear regression model was developed to predict whole blood volume (Fig. 2b). The mass of unused psDBS membranes (HemaSpot) was measured to assess baseline variability (mean ± SD: 31.2 ± 0.62 mg, CV: 2.0%), confirming low device-to-device variation. The model was trained on a group of psDBS (HemaSpot) samples pipetted at 50 to 250 µL in the Murtaza lab using whole blood samples obtained from healthy individuals (enrolled at the UWCCC BioBank). The model was tested on an independent group of samples collected and pipetted at 150 µL by UWCCC BioBank staff using whole blood samples from additional healthy individuals and patients with cancer (Table 1). Pearson’s r and root mean squared error (RMSE) were used to assess model accuracy.

To evaluate the generalizability of the model across psDBS collection devices, the model was trained on a group of psDBS samples pipetted at 30 to 70 µL in the Murtaza lab onto HemaSep Strips, using whole blood samples obtained from healthy individuals enrolled at the UWCCC BioBank. This model was then tested on an independent group of samples from healthy individuals and patients with cancer and pipetted at 50 µL either by UWCCC BioBank staff or members of the Murtaza lab. Pearson’s r and root mean squared error (RMSE) were used to assess model accuracy.

### Circulating plasma DNA extraction and quantification

DNA was extracted with the MagMAX Cell-Free DNA Isolation Kit (Applied Biosystems, Foster City, CA) using a modified protocol adapted for psDBS samples as described earlier [5]. Extracted DNA was assessed and quantified using a real-time PCR (qPCR) assay targeting the L1PA2 gene on the QuantStudio 6 Pro system (Applied Biosystems, Foster City, CA). A 10 to 20 μL reaction of the Luna[1] Universal qPCR Master Mix kit was performed following the manufacturer’s instructions. The primer sequences used were from Breitbach *et al*. and are listed in supplemental table S1 [10]. All templates were measured in duplicate or triplicate. All reactions comprised a triplicate of non-template controls (NTC).

Amplification consisted of an initial denaturation at 95°C for 60 seconds, followed by 40 cycles of denaturation at 95°C for 15 seconds, and extension at 60°C for 30 seconds. Amplification finished with a melt curve starting at 95°C for 1 second, 60°C for 20 seconds, and 95°C for 1 second. Subsequent qPCR measurements were calibrated to a standard dilution series.

## Results

### Correlation of extracted features with known blood volumes

To evaluate whether there is a correlation between mass, area, and brightness and the actual volume of blood spotted on the psDBS, we first applied simple linear regression for each of these variables.

We analyzed a set of 250 psDBS samples (Hemaspot) prepared using commercially obtained whole blood from 30 individuals with total spotted blood volumes ranging from 50 to 250 µL. The strongest linear correlation was observed between volume and mass (Pearson r = 0.95, RMSE = 4.2 mg), followed by volume and area (Pearson r = 0.87, RMSE = 0.52 cm²; Fig. 1b). We also observed a negative correlation between volume and MGV (Pearson r = -0.69, RMSE = 7.9), consistent with our hypothesis that a higher volume is correlated with a dark-colored sample (Fig. 1b).

### Accuracy and performance of the volume inference model

To predict blood volume collected on psDBS, we developed a multiple linear regression model using mass and imaging features: mass of the whole psDBS, area of the erythrocyte region, and brightness of the erythrocyte region (supplemental table S2). Using 110 samples from 10 healthy individuals (model training dataset), a linear regression model was fit to the data, showing a strong correlation between actual and predicted blood volume (Pearson r = 0.97, RMSE = 5.1 μL; Fig. 3a-b). Using an independent cohort of 151 samples from 33 separate healthy individuals and 118 patients with cancer (model testing dataset), the model prediction had minimal deviations from the actual volume (RMSE = 13.7 μL), although it slightly underestimated the true volume, with a median error of -5.8% (Fig. 3c). When separating the model testing dataset between healthy individuals and patients with cancer, we found comparable performance and a similar difference between the actual volume and the predicted volume in both sets of samples (healthy median error = -6.5%, cancer = - 5.5%; Fig. 3d-e).

**Fig. 3:**
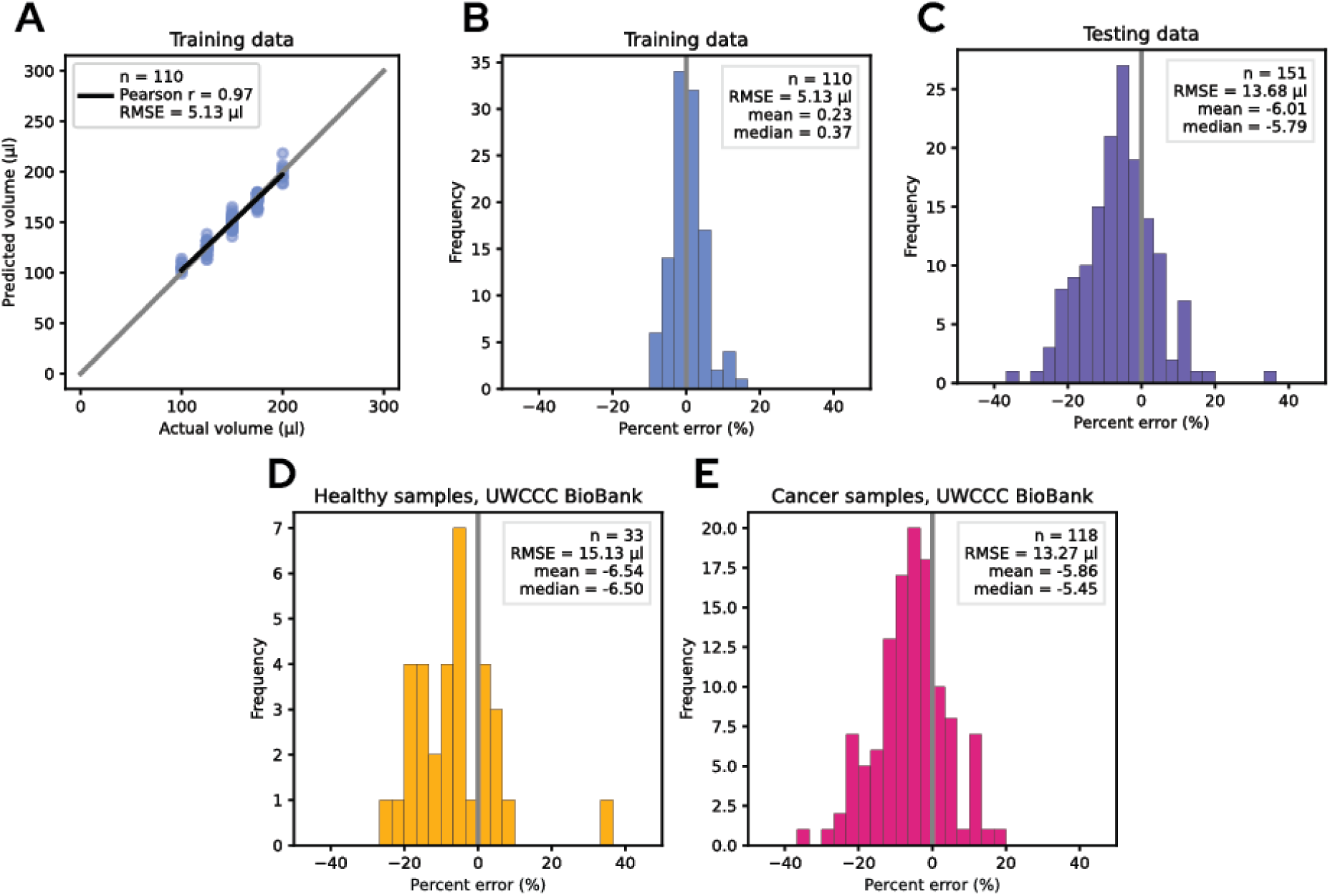
Training and evaluation of a linear regression model to infer blood volume collected on HemaSpot samples. **(a)** The model was trained using 110 samples from 10 healthy individuals. A strong correlation between actual and predicted blood volume was observed (Pearson r = 0.97), with a strong predictive performance (RMSE = 5.1 μL) across a measured range of 100 to 200 μL. **(b)** Using the same dataset as in Fig. 3a, the percent error between predicted and actual blood volumes was centered near zero (mean = 0.23%, median = 0.37%). **(c)** The model was tested using 151 samples from 33 healthy individuals and 118 patients with cancer. All samples were 150 μL. Remarkable predictive performance between actual and predicted blood volume was observed (RMSE = 13.7 μL), with a slight underestimation bias (mean = -6.0%, median = -5.8%). **(d)** Using 33 samples from 33 healthy individuals from the same dataset as in Fig. 3c, similar predictive performance between actual and predicted blood volume was observed (RMSE = 15.1 μL), with a slight underestimation bias (mean = -6.5%, median = -6.5%). **(e)** Using 118 samples from 118 patients with cancer from the same dataset as in Fig. 3c, similar predictive performance between actual and predicted blood volume was observed (RMSE = 13.3 μL), with a slight underestimation bias (mean = -5.9%, median = -5.5%).

### Application of the inference approach to HemaSep Strip device

To evaluate the generalizability of the volume inference approach across psDBS devices, we re-trained and evaluated the inference model to samples collected using the HemaSep Strip psDBS collection device. We collected a total of 80 samples from 20 healthy individuals and 20 patients with cancer, using volumes ranging from 30 µL to 70 µL per strip (supplemental table S3). Despite differences in device profile, total blood volume, and sample spreading characteristics, we observed consistent relationships between blood volume and extracted features (fig. S2). Using 50 samples from 10 healthy individuals, a linear regression model was fit to the data, showing a strong correlation between actual and predicted blood volume (Pearson r = 0.97) and a strong predictive performance (RMSE = 2.6 μL; Fig. 4a-b, supplemental table S4). Using 30 samples from 10 separate healthy individuals and 20 patients with cancer to test the model, the model prediction had minimal deviations from the actual volume (RMSE = 6.7 μL), although it slightly overestimated the true volume, with a median error of 4.4% (Fig. 4c).

**Fig. 4:**
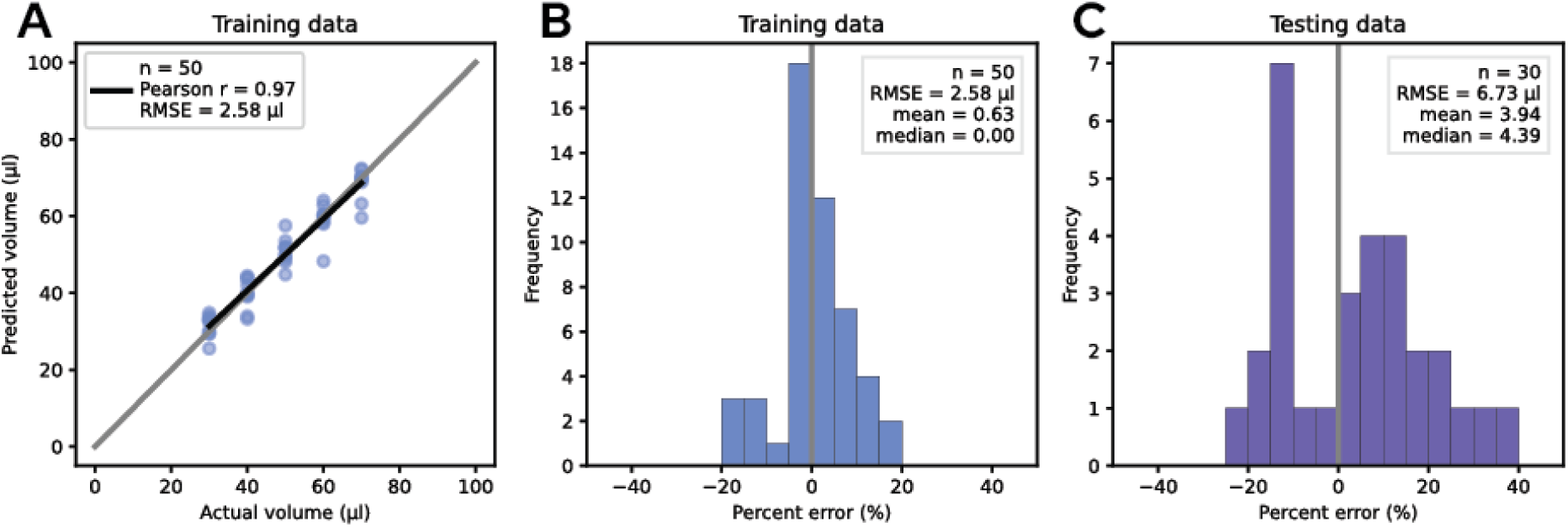
Training and evaluate of a linear regression model to infer blood volume collected on HemaSep strips. **(a)** The model was trained using 50 samples from 10 healthy individuals. A strong correlation between actual and predicted blood volume was observed (Pearson r = 0.97), with a strong predictive performance (RMSE = 2.6 μL) across a measured range of 30 to 70 μL. **(b)** Using the same dataset as in Fig. 4a, the percent error between predicted and actual blood volumes was centered near zero (mean = 0.63%, median = 0.00%). **(c)** The model was tested using 30 samples from 10 healthy individuals and 20 patients with cancer. All samples were 50 μL. Remarkable predictive performance between actual and predicted blood volume was observed (RMSE = 6.7 μL), with a slight overestimation bias (mean = 3.9%, median = 4.4%).

### Correlation of volume-adjusted psDBS and matched plasma cfDNA

After DNA extraction and quantification of cfDNA in eluted samples using qPCR, we calculated circulating cfDNA concentrations in psDBS using inferred blood volumes. We compared these measurements with plasma cfDNA concentrations in matched blood tubes (measured using automated gel electrophoresis). Plasma cfDNA concentrations are represented as ng cfDNA per mL of plasma, while psDBS cfDNA concentrations are represented as ng cfDNA per mL of whole blood.

To confirm concordance between qPCR and gel electrophoresis, we initially compared DNA concentrations using the two quantification methods in the same set of plasma samples. We observed comparable cfDNA concentration estimations (R² = 0.91, slope = 1.10; fig. S3). cfDNA concentrations measured in psDBS and plasma samples were not normally distributed. Using inferred blood volumes from psDBS to calculate circulating concentrations, we observed a moderate correlation between psDBS cfDNA and matched plasma cfDNA concentrations (Spearman’s rho = 0.62; Fig. 5).

**Fig. 5.**
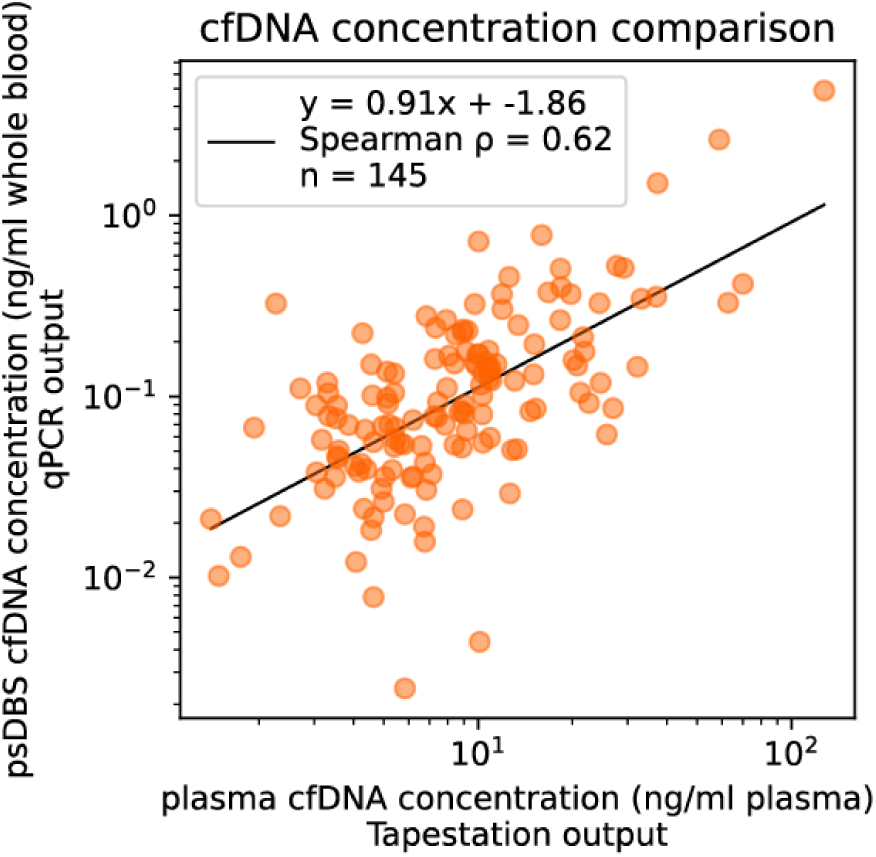
Comparison of circulating plasma DNA concentration measured from blood tubes and psDBS (HemaSpot) samples. Inferred blood volumes in psDBS samples were used to calculate circulating plasma DNA concentrations and compared with corresponding matched blood tubes. Two samples were removed after failed quantification. Both axes are log transformed to accommodate skewed plasma cfDNA distributions, and moderate correlation was observed.

## Discussion

Plasma-separating dried blood spots may be useful as a low-cost, logistically simple alternative to collection, processing, shipping and storage of blood tubes that could improve access to biomarkers for cancer detection and monitoring in resource-limited environments. However, unlike phlebotomy to collect blood tubes where the volume of blood or plasma collected and analyzed can be directly measured, volume of blood collected in psDBS samples is not directly measured and can vary based on user experience, collection method, physiological factors as well as the collection environment. In the current study, we observed that blood volume collected in psDBS samples can be inferred with remarkable accuracy using mass and image analysis. We further observed that circulating concentration of plasma DNA calculated using the inferred blood volume was correlated with plasma DNA concentration in corresponding blood tubes.

Previous studies have used imaging and gravimetric analysis to estimate blood hematocrit or infer blood volume in DBS [12], [15], [16], [13], [14]. However, these approaches were developed for conventional dried whole blood spots. In contrast to whole blood spots, the spreading of blood during separation on plasma membranes yields additional image features that can potentially be combined to overcome prior limitations. In particular, hematocrit or the proportion of total blood volume contributed by erythrocytes can affect density, color and spread of blood DBS [9]. Hematocrit levels are generally higher in men compared to women, though factors like age and hydration can shift these values[11]. Although previous studies have found hematocrit levels to be a significant confounder[11], [12], [13], [14], our validation results across 151 individuals showed good accuracy and no biases between males and females. This is likely due to our combined analysis of mass, color intensity and area of blood spread, features that can be unique measured from psDBS and not whole blood spots.

There are several limitations of the current study that require further work. First, we observed a systematic underestimation of inferred blood volumes in our validation dataset by about 6%. One potential explanation is that while samples for the training dataset were prepared by one set of operators, the validation set was prepared independently by a different set of operators, representing differences in lab environment, equipment and sample handling. Another potential explanation is that all validation samples were prepared at 150 μL blood volume, and the predictive value and bias of mass, color intensity and area of blood spread may vary at different ranges of blood volume collected, though we don’t see clear evidence of this in the training dataset. Finally, it is possible that these differences are driven by physiological differences between the training cohort which was only comprised of healthy individuals and patients with cancer comprised most of the validation cohort. Second, our current validation cohort is comprised of prepared blood spots where a known volume of blood was pipetted on psDBS membrane. This is required to estimate error in predicted volumes but does not directly mimic the real world scenario where fingerstick samples must be evaluated to infer collected blood volume. These limitations can be potentially addressed by future studies with larger sample sizes and real world psDBS collections.

We further note that although cfDNA concentrations observed in psDBS are correlated with corresponding blood samples, cfDNA concentration in psDBS are lower than expected even after accounting for collected blood volume (Fig. 5). While its currently unclear what drives this discrepancy, potential drivers included limited efficiency of the cfDNA extraction protocol we have used, degradation of DNA during plasma separation or psDBS storage, or differences in separation of extracellular DNA from cellular components on a plasma membrane compared to centrifugation. These factors and the impact of different matrices used for psDBS need further evaluation to improve cfDNA yield from psDBS. We also observed challenges associated with precisely and accurately quantifying cfDNA at such low concentrations. Although the method we used here targets a repetitive locus on the human genome to improve quantification[10], the lowest measurable standard in our assay corresponds to approximately 12 targets of the L1PA2 element, limiting the precision due to the large impact of sampling variation at such concentrations.

We found that mass, area of blood spread and intensity were informative of blood volume collected using two different psDBS devices, with distinct profiles and volume ranges. However, for each device, we adapted our image segmentation approach and re-trained the model. Hence, differences in psDBS devices such as profile, membrane material, porosity, separation efficiency and device-to-device variation mean that a model trained on one psDBS device will not directly apply to another. At minimum, such models may need to be re-trained and may require additional feasibility studies for each new device used.

In summary, our work demonstrates that it is feasible to estimate blood volume and cfDNA concentrations with remarkable precision and accuracy in psDBS samples by combining gravimetric and imaging features, without the need to explicitly account for hematocrit. This approach can enable calculation of circulating concentrations for serum and plasma biomarkers measured from psDBS across different disease types including for patients with cancer.

## Data Availability

All data produced in the present study are available upon reasonable request to the authors.

## Acknowledgements

Thank you to Mr. Will Suter, Ms. Hellen Nginga, and the UW Carbone Cancer Center BioBank for sample collection and processing. Supported by the UW Carbone Cancer Center Global Oncology Grant program and NIH grants U01CA243078, R01CA223481, and P30CA014520. Thank you to current and former members of the Murtaza Lab for guidance and support. Some figures were created with BioRender.com.

## Statement of translational relevance

Serum and plasma biomarkers are widely used for cancer diagnostics, detection, and monitoring. Recent development of plasma-separating dried blood spots (psDBS) can enable a low-cost, logistically simple alternative to phlebotomy for at-home and distributed sample collection, as well as storage and shipping at room temperature. However, to calculate the circulating concentration of any biomarker, the original volume of sample collected in a blood spot is required, and most psDBS devices are non-volumetric. In this study, we developed an approach to infer blood volume collected on plasma-separating DBS (psDBS) using its mass and scanned image. Using inferred volumes, we estimated concentration of plasma DNA as an example in a group of healthy individuals and patients with cancer. This approach can enable the use of psDBS to improve access for blood-based cancer biomarker testing in rural areas with limited healthcare resources.

## Supplemental material

**Fig. S1.**
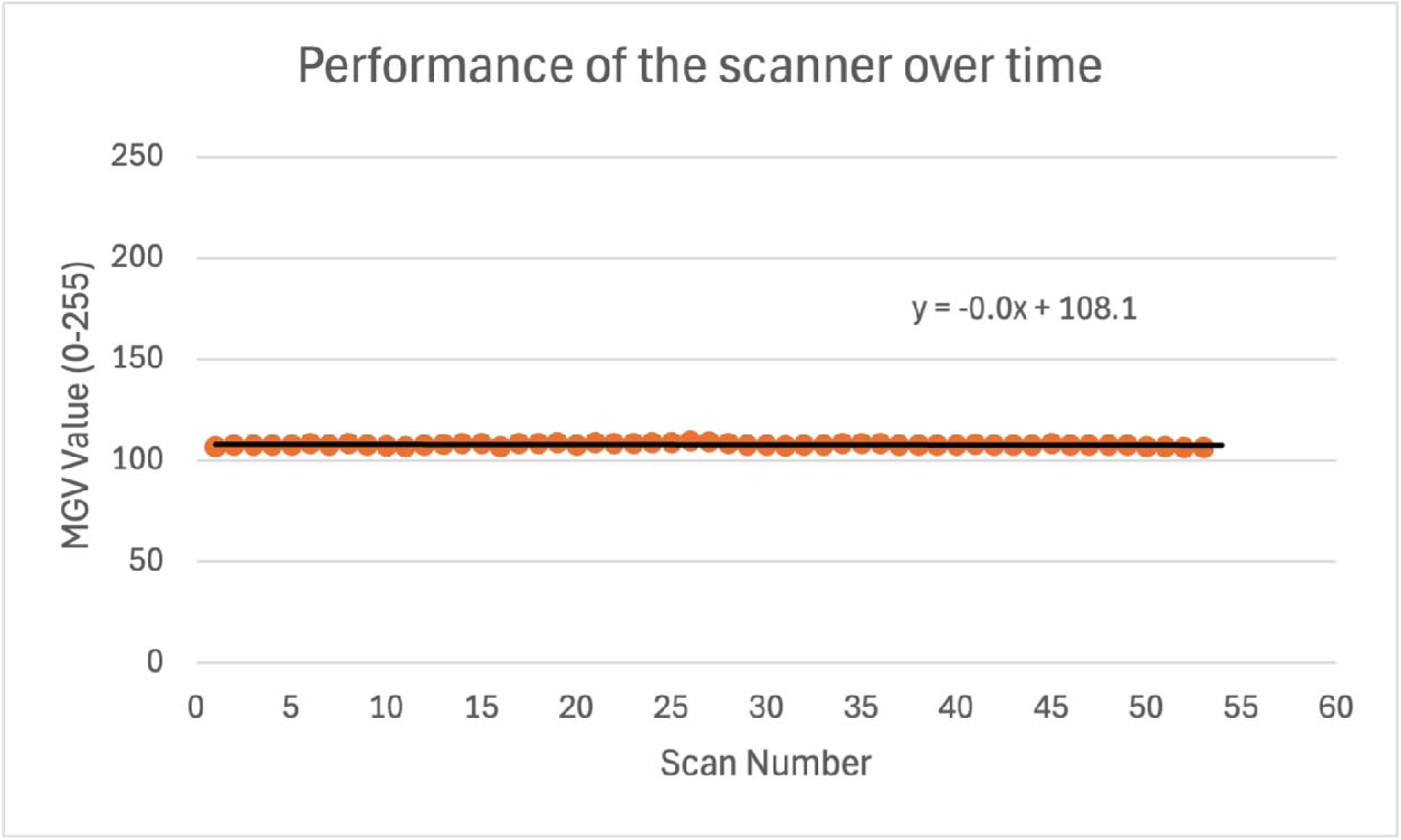
To ensure scanner performance, a Delta 1 gray card (B&H Photo, New York, NY) was scanned before, during, and/or after scanning any psDBS. The MGV ranges from 0-255 on a unitless scale, and the scan number corresponds to the order the greycard scans were collected, and represents the scans taken over the entire collection period.

**Supplemental table S1.**
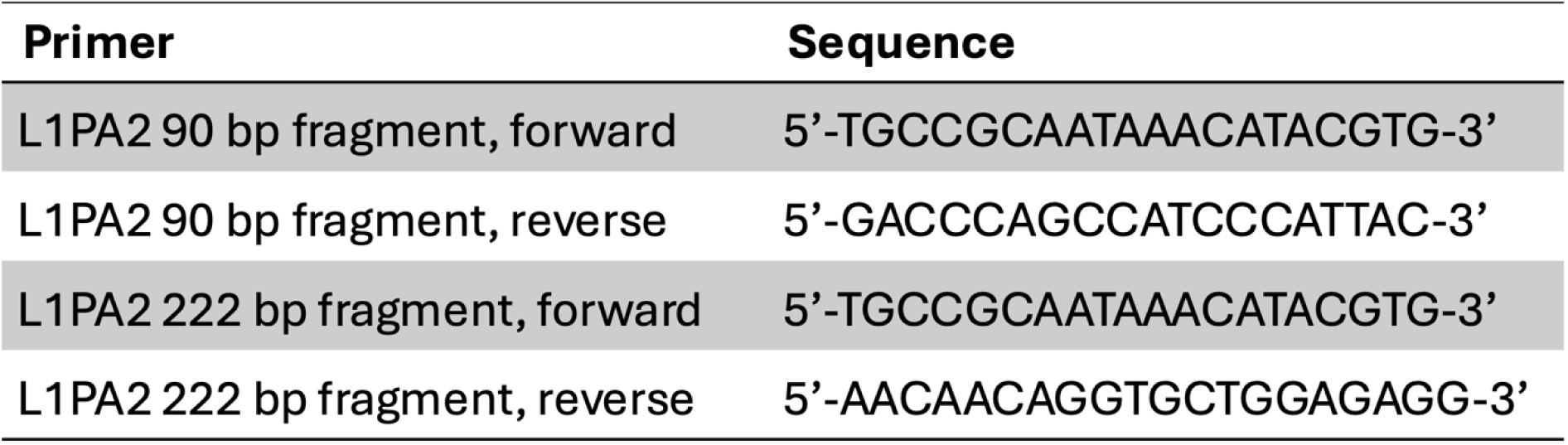
L1PA2 primer sequences for the qPCR assay.

**Supplemental table S2.** OLS regression results for Hemaspot psDBS samples.

| Predictor | $\beta$ (SE) | t | p | 95% CI |
| --- | --- | --- | --- | --- |
| Intercept | 150.00 (0.63) | 239.36 | <0.001 | [148.76, 151.24] |
| Net mass (z-score) | 20.87 (1.98) | 10.53 | <0.001 | [16.94, 24.80] |
| MGV (z-score) | -3.82 (1.42) | -2.69 | 0.008 | [-6.64, -1.00] |
| Area (z-score) | 4.42 (1.21) | 3.65 | <0.001 | [2.02, 6.81] |

$R^2 = 0.948$
Adjusted $R^2 = 0.946$
F (3, 106) = 639.8
p < 0.001

**Supplemental table S3.** HemaSep Strip cohort table.

|  | Source | Samples<br>n (%) | Individuals<br>n (%) | Age<br>median (range) | Sex<br>n (%) | Disease Status<br>n (%) |
| --- | --- | --- | --- | --- | --- | --- |
| Feature exploration & model training | UWCCC Biobank | 50 (100) | 10 (100) | NA | F: 7 (70)<br>M: 3 (30) | Healthy: 10 (100) |
| Model testing | UWCCC Biobank | 30 (100) | 30 (100) | NA | M: 11 (36.7)<br>F: 19 (63.3) | Healthy: 10 (33.3)<br>Cancer: 20 (66.7) |

**Fig. S2.**
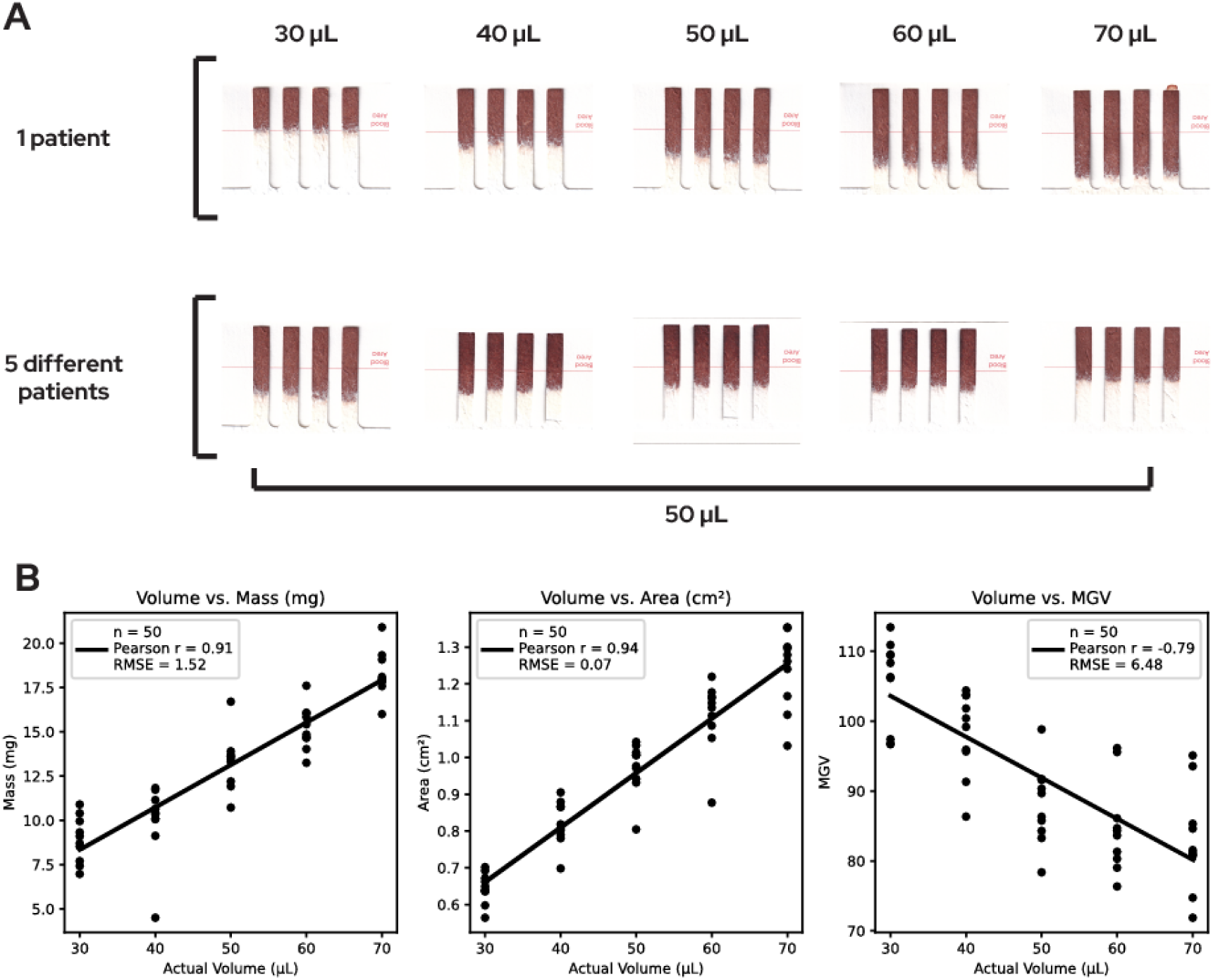
In the HemaSep Strip, samples of different volumes from one patient have visible differences in the erythrocyte area and the brightness of the psDBS as volume increases. Samples of the same volume from five different patients have visible differences in the erythrocyte area and the brightness of the DBS due to biological variability. **Fig. S2b.** There are correlations between the actual volume of the sample and mass and area (Pearson r = 0.91 and Pearson r = 0.94, respectively). There is a negative correlation between the actual volume of the sample and MGV (Pearson r = -0.79).

**Supplemental table S4.**
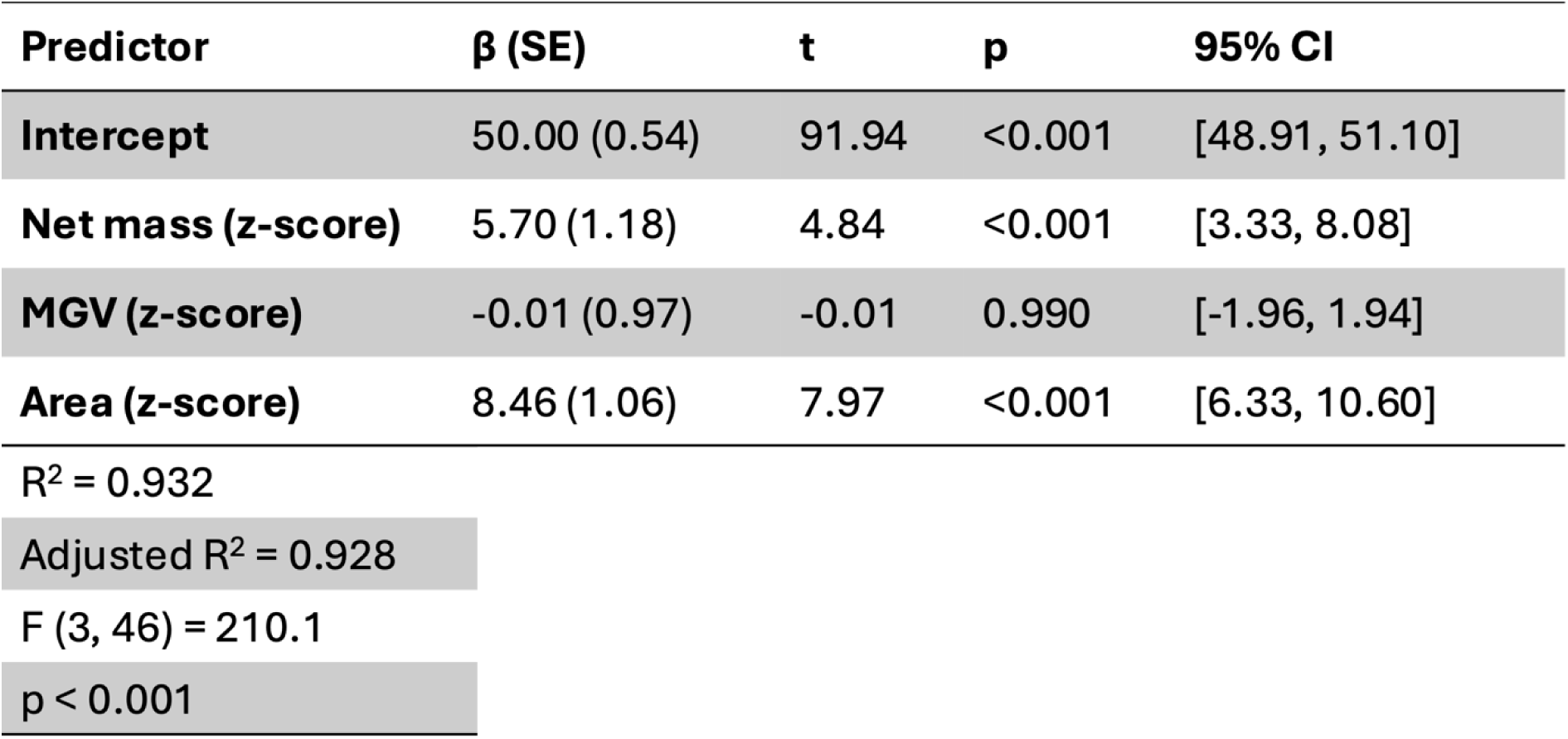
HemaSep Strip device OLS regression results.

| Predictor | $\beta$ (SE) | t | p | 95% CI |
| --- | --- | --- | --- | --- |
| Intercept | 50.00 (0.54) | 91.94 | <0.001 | [48.91, 51.10] |
| Net mass (z-score) | 5.70 (1.18) | 4.84 | <0.001 | [3.33, 8.08] |
| MGV (z-score) | -0.01 (0.97) | -0.01 | 0.990 | [-1.96, 1.94] |
| Area (z-score) | 8.46 (1.06) | 7.97 | <0.001 | [6.33, 10.60] |

$R^2 = 0.932$
Adjusted $R^2 = 0.928$
F (3, 46) = 210.1
p < 0.001

**Fig. S3.**
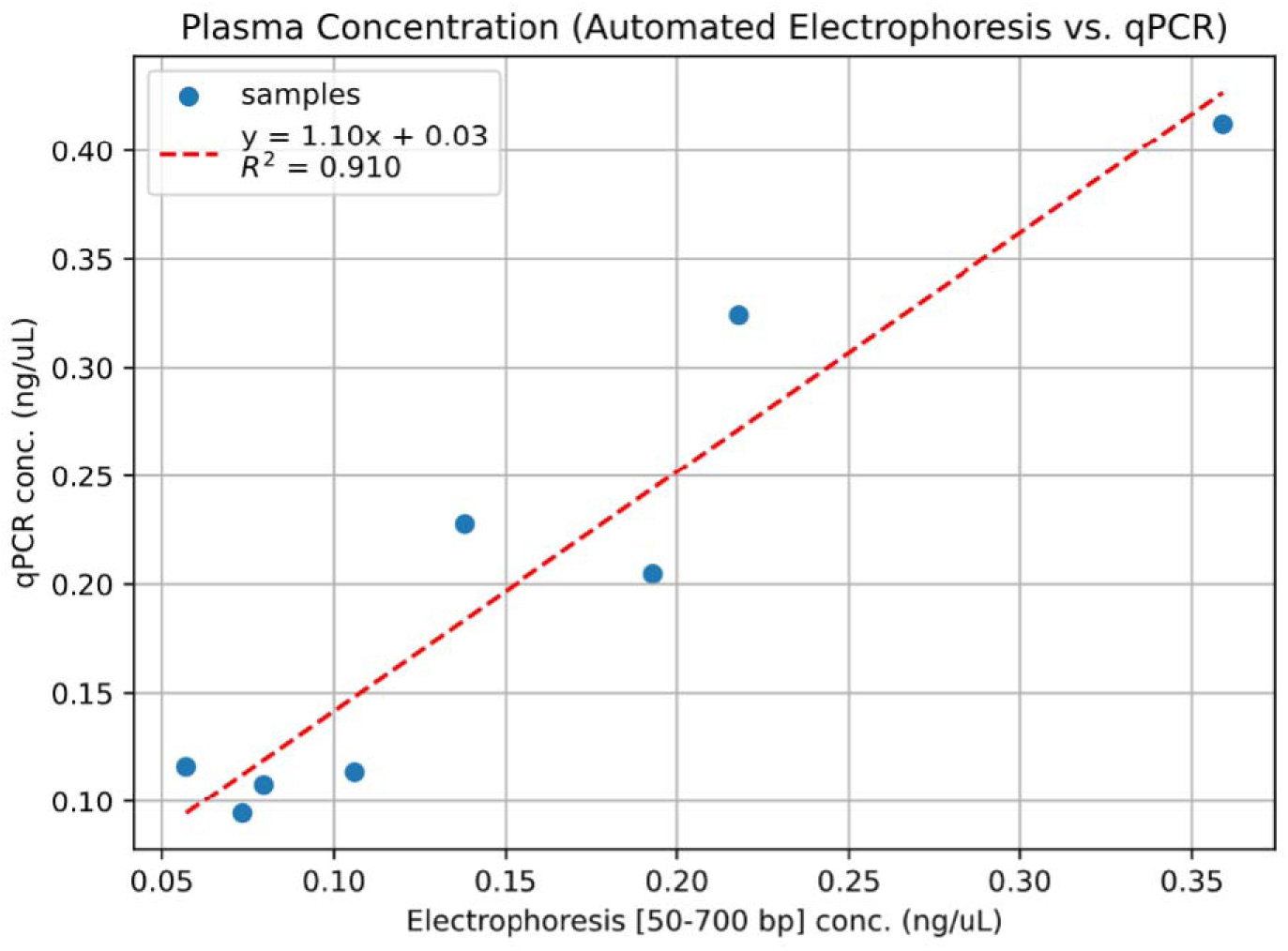
To confirm assay concordance, we compared DNA concentrations using two quantification methods, an automated gel electrophoresis method and the L1PA2 qPCR assay using 8 samples. We observed comparable cfDNA concentration estimations with an R² = 0.91 and a slope = 1.10.

